# Genomic epidemiology of SARS-CoV-2 in Guangdong Province, China

**DOI:** 10.1101/2020.04.01.20047076

**Authors:** Jing Lu, Louis du Plessis, Zhe Liu, Verity Hill, Min Kang, Huifang Lin, Jiufeng Sun, Sarah François, Moritz U G Kraemer, Nuno R Faria, John T McCrone, Jingju Peng, Qianlin Xiong, Runyu Yuan, Lilian Zeng, Pingping Zhou, Chuming Liang, Lina Yi, Jun Liu, Jianpeng Xiao, Jianxiong Hu, Tao Liu, Wenjun Ma, Wei Li, Juan Su, Huanying Zheng, Bo Peng, Shisong Fang, Wenzhe Su, Kuibiao Li, Ruilin Sun, Ru bai, Xi Tang, Minfeng Liang, Josh Quick, Tie Song, Andrew Rambaut, Nick Loman, Jayna Raghwani, Oliver G Pybus, Changwen Ke

## Abstract

COVID-19 is caused by the SARS-CoV-2 coronavirus and was first reported in central China in December 2019. Extensive molecular surveillance in Guangdong, China’s most populous province, during early 2020 resulted in 1,388 reported RNA positive cases from 1.6 million tests. In order to understand the molecular epidemiology and genetic diversity of SARS-CoV-2 in China we generated 53 genomes from infected individuals in Guangdong using a combination of metagenomic sequencing and tiling amplicon approaches. Combined epidemiological and phylogenetic analyses indicate multiple independent introductions to Guangdong, although phylogenetic clustering is uncertain due to low virus genetic variation early in the pandemic. Our results illustrate how the timing, size and duration of putative local transmission chains were constrained by national travel restrictions and by the province’s large-scale intensive surveillance and intervention measures. Despite these successes, COVID-19 surveillance in Guangdong is still required as the number of cases imported from other countries is increasing.

**Highlights:**

- 1.6 million molecular diagnostic tests identified 1,388 SARS-CoV-2 infections in Guangdong Province, China, by 19^th^ March 2020
- Virus genomes can be recovered using a variety of sequencing approaches from a range of patient samples.
- Genomic analyses reveal multiple virus importations into Guangdong Province, resulting in genetically distinct clusters that require careful interpretation.
- Large-scale epidemiological surveillance and intervention measures were effective in interrupting community transmission in Guangdong

---

A new virus-associated disease, COVID-19 (coronavirus disease 2019), was initially reported in China on 30^th^ December 2019 (Wu et al., 2020). The causative agent of COVID-19 is the novel human coronavirus SARS-CoV-2 (Wu et al., 2020; Zhou et al., 2020), and as of 24^th^ March 2020, there have been 372,757 confirmed infections and 16,231 deaths reported worldwide (WHO, 2020). In China, the COVID-19 epidemic grew exponentially during January 2020, peaking on 12^th^ February 2020 with 15,153 newly confirmed cases per day. One month later, reported COVID-19 cases in China dropped to approximately 20 per day, suggesting the epidemic there was contained. However, since the second half of February 2020, the number of cases reported outside of China has risen exponentially. By 11^th^ March 2020, the day that WHO announced COVID-19 as a new pandemic, 37,371 cases had been reported outside of China (WHO, 2020).

Guangdong Province and the Pearl River Delta Metropolitan Region contain some of the world’s largest and most densely populated urban areas. Guangdong is the most populous province of China (113m people) and contains many large cities including Guangzhou (12m), Shenzhen (10m), Dongguan (8m) and Foshan (7m). The province has strong transportation links to Hubei Province, where the first cases of COVID-19 were reported. The Wuhan–Guangzhou high-speed railway has been estimated to transfer 0.1–0.2 million passengers per day during the spring festival period, which started on 10^th^ January 2020. By 19^th^ March 2020, Guangdong had 1,388 confirmed cases of COVID-19, the highest in China outside of Hubei Province.

Understanding the evolution and transmission patterns of a virus after it enters a new population is crucial for designing effective strategies for disease control and prevention (Grubaugh et al., 2017; Ladner et al., 2019). In this study, we combine genetic and epidemiological data to investigate the genetic diversity, evolution, and epidemiology of SARS-CoV-2 in Guangdong Province. We generated virus genome sequences from 53 patients in Guangdong and sought to investigate the timing and relative contributions of imported cases versus local transmission, the nature of genetically-distinct of transmission chains within Guangdong, and how the emergency response in Guangdong was reflected in the reduction and elimination of these transmission chains. Our data may provide valuable information for implementing and interpreting genomic surveillance of COVID-19 in other regions.

Enhanced surveillance was launched in all clinics in Guangdong province following the first reports of patients with undiagnosed pneumonia on 30^th^ December 2019. Initially, screening and sampling for SARS-CoV-2 was targeted towards patients with fever and respiratory symptoms and who had a history of travel in the 14 days before the date of symptom onset. The first detected case had symptom onset on 1^st^ January and was reported on 19^th^ January 2020 (Kang et al. 2020). COVID-19 cases in Guangdong grew until early February 2020 (peaking at >100 cases per day) and declined thereafter (Figure 1A). After 22^nd^ February 2020, the daily number of locally-infected reported cases in Guangdong did not exceed one. However, since the beginning of March 2020, COVID-19 cases imported to Guangdong from abroad have been detected with increasing frequency. As of 26^th^ March 2020, a total of 102 imported cases were reported from 19 different countries (Figure 1A), highlighting the risk that local COVID-19 transmission could reignite in China.

**Figure 1:**
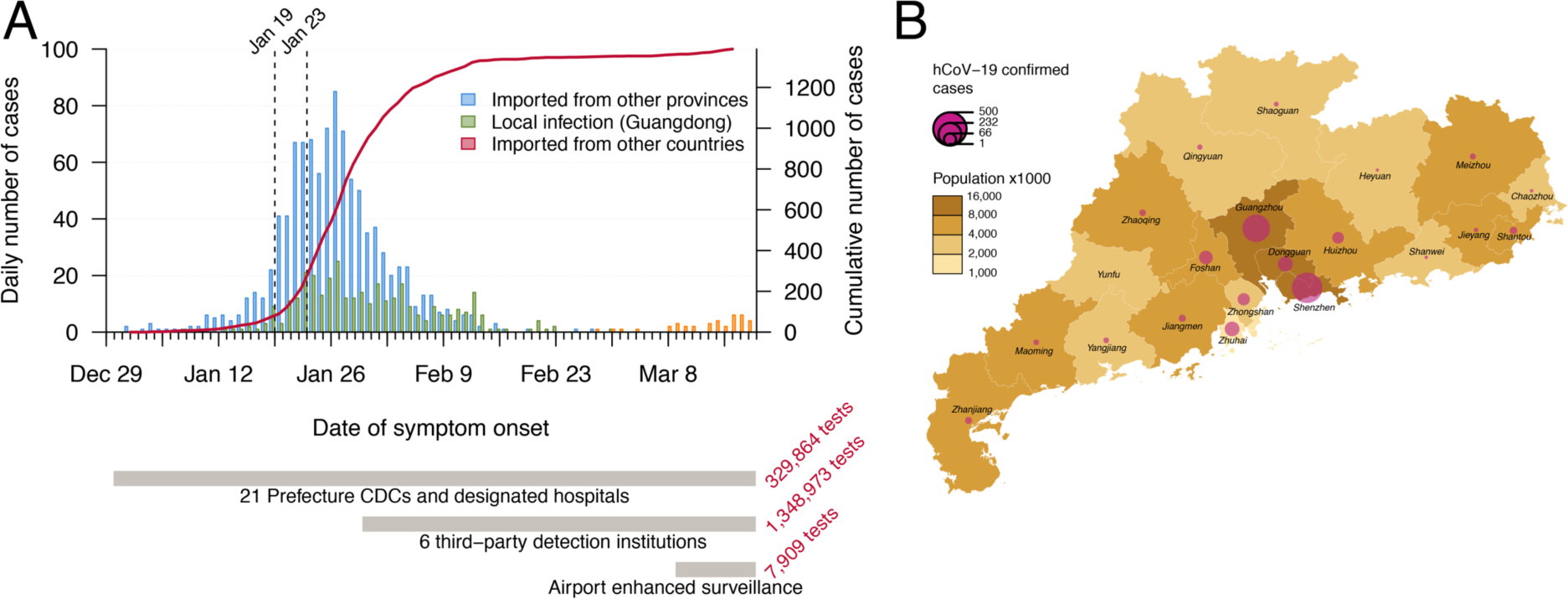
Summary of the COVID-19 epidemic in Guangdong Province, China. (A) Time series of the 1,388 laboratory-confirmed COVID-19 cases in Guangdong until 19^th^ March, by date of onset of illness. Cases are classified according to their likely exposure histories (see inset and main text). The dashed lines indicate the date the first Guangdong case was detected (19^th^ January) and the shutdown of travel from Wuhan (23^rd^ January). An overview of testing and surveillance strategies at different stages of the epidemic are illustrated below the time series, on the same timescale. (B) Geographic distribution of COVID-19 cases and human population density among the 21 prefecture-level divisions of Guangdong Province.

Different surveillance strategies were applied during the epidemic in Guangdong (Figure 1A). More intense surveillance was initiated on 30^th^ January 2020 in response to the Spring Festival period, which results in greater mobility among regions and provinces in China (Kraemer, 2020) and because asymptomatic COVID-19 cases had been reported (Guan et al., 2020). This included monitoring (i) all travelers returning from Hubei or other regions with high epidemic activity, (ii) their close contacts, and (iii) all hospitalized patients in clinics, including those without fever or respiratory symptoms, regardless of their exposure history. Approximately 1.35 million samples were screened by six third-party institutions between 30^th^ January and 15^th^ March 2020. Surveillance commenced at Guangdong airports in early March, following the growth of COVID-19 outbreaks outside of China. In total, approximately 1.6 million tests were performed by 19^th^ March, identifying 1,388 SARS-CoV-2 positive cases in 20 of 21 prefectures in Guangdong Province (Figure 1B). Around a quarter of cases (336) were judged to be linked to local transmission and two-thirds (1014) had a likely exposure history in Hubei (see Methods). For locally-infected cases, 181 (53%) were linked to transmissions among household members. More than half of reported cases (60%) were from the cities of Shenzhen and Guangzhou (Figure 1B). We note that the number of detected cases will be less than the true number of infections, although the degree of under-reporting is unknown. Surveillance was targeted towards travellers, hence these data may overestimate the proportion of travel-associated cases.

To understand the genetic structure of the COVID-19 outbreak in Guangdong, we generated near-complete and partial genomes from 53 COVID-19 patients in Guangdong Province. The genomes were generated by a combination of metagenomic sequencing and multiplex PCR amplification followed by nanopore sequencing on a MinION device (see Methods). Sequence sampling dates ranged from 30^th^ January to 28^th^ February 2020 (Figure S1).

Sequencing was performed on 79 clinical samples (throat swabs, n=32, anal swabs n=24, nasopharyngeal swabs n=10, sputum n=13) collected from 65 patients with varying disease symptoms, ranging from asymptomatic to very severe (see methods). RT-PCR Ct (cycle threshold) values of these samples ranged from 19 to 40.86. Figure 2A displays the Ct values for the 53 samples with >50% genome coverage for which we report whole and partial genome sequences (see Figure S2 for details of all 79 samples). When Ct values are <30, sequence reads covered approximately 90% or more of the reference genome (GenBank accession number: *MN908947*.*3*) irrespective of the amplification and sequencing approach used (see Methods). However, genome coverage declined for samples with Ct >30 (Figure S2). Using a Kruskal-Wallis rank sum test we found an association between sample Ct values and sample type (Figure 2B; *p*<0.001), and sample Ct values and disease severity (Figure 2C; *p*=0.03; see also Liu et al. 2020).

**Figure 2:**
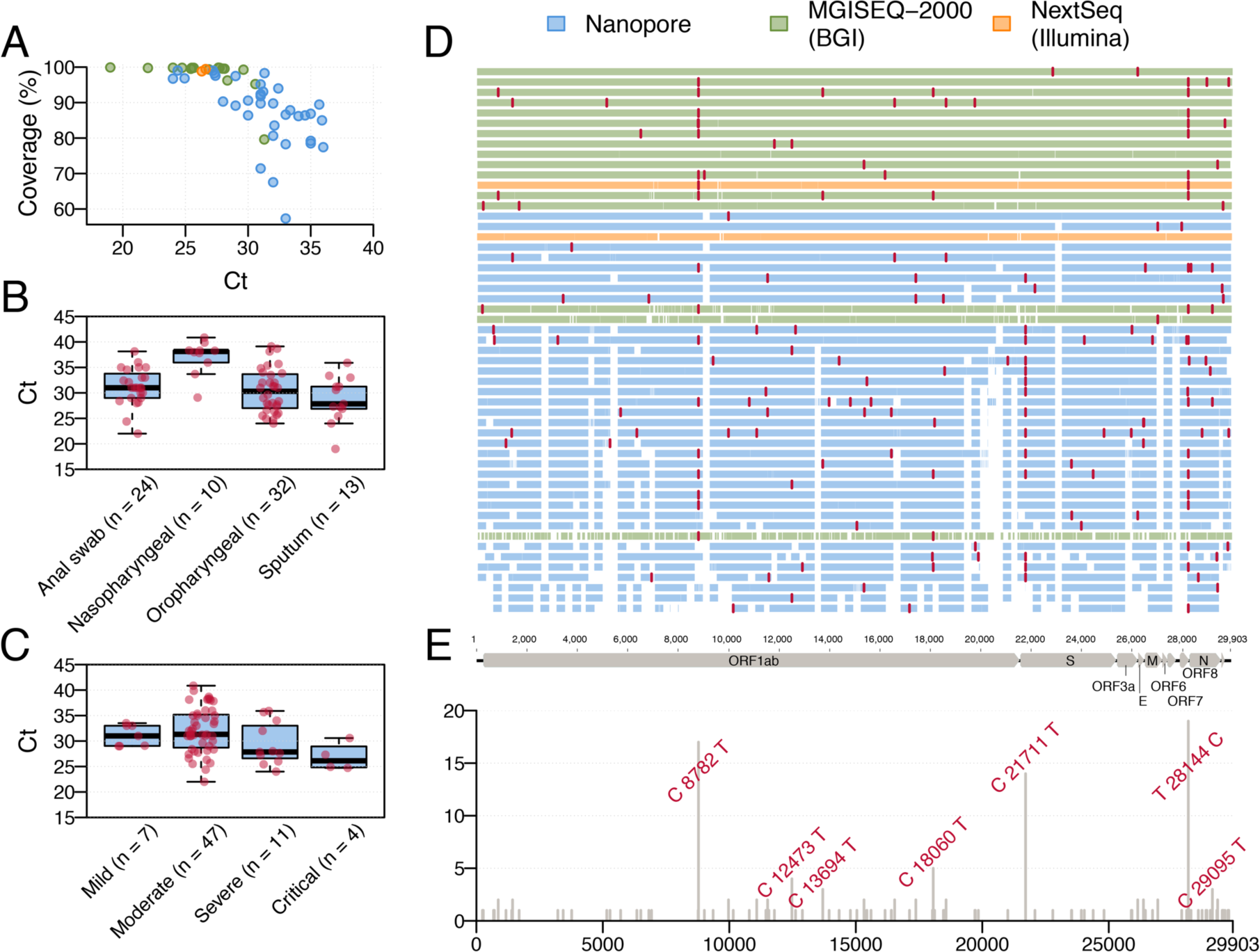
(A) Plot of SARS-CoV-2 genome coverage against RT-PCR Ct value for the 53 genomes reported here. Each sequence is coloured by sequencing approach: blue = BGI metagenomic sequencing, orange = multiplex PCR nanopore sequencing, green = NextSeq metagenomic sequencing. (B) RT-PCR Ct values for different sample types. (C) RT-PCR Ct values for samples from patients with different disease severity; the ‘mild’ category includes 2 asymptomatic cases. (D) Genome coverage map for the 53 genomes reported here, ordered by coverage. Single nucleotide variants (with respect to the reference genome MN908947.3) are coloured in red. Each genome (row) is coloured according to sequencing approach. (E) Genomic structure of SARS-CoV-2 and the genomic location and frequency of single nucleotide variants (with respect to the reference genome MN908947.3) among our 53 sequences. These variants are also shown as red lines in panel D.

Sequences generated with nanopore sequencing indicate common regions of low coverage (Figure 2D). The version 1 primer set used here was not able to amplify some regions with high frequency of changes from the reference genome *MN908947*.*3*. After completion of this study, the primers have been redesigned to improve coverage (Quick 2020). Shared and unique single nucleotide variants (SNVs) were observed at 97 sites across the virus genomes (Figure 2D, 2E), with 77 SNVs present in only one genome. Three SNVs were present in >10 genomes: (C8782T, C21711T and T28144C). When compared to 49 previously released genomes from Hubei and Guangdong, 118 SNVs are present in only one genome and the same three SNVs are still the only SNVs shared between more than 10 genomes (Figure S3).

To understand the genetic diversity of the SARS-CoV-2 epidemic in Guangdong we performed phylogenetic analyses using maximum likelihood and Bayesian molecular clock approaches. We added our new virus genomes from Guangdong to 177 publicly-available sequences, which includes 73 sequences from China, 17 of which are previously-reported Guangdong genomes. The final alignment comprised 250 sequences and increased the number of SARS-CoV-2 sequences from China by ∼60% when our data was submitted to GISAID (on 9^th^ March).

The estimated maximum likelihood (ML) phylogeny is shown in Figure 3A. The SARS-CoV-2 sequences from Guangdong (red) are interspersed with viral lineages sampled from other Chinese provinces and other countries (black). This pattern agrees with the time series in Figure 1A, indicating that most detected cases were linked to travel rather than local community transmission. Despite this, there were a number of instances where sequences from Guangdong appeared to cluster together, sometimes with sequences sampled from other regions. To explore these lineages in more detail, we performed a Bayesian molecular clock analysis that places the phylogenetic history of the genomes on an estimated timescale. A summary visualisation of the maximum clade credibility tree from that analysis is shown in Figure 3B and is largely congruent with the ML tree. The current low genetic diversity of SARS-CoV-2 genomes worldwide means that most internal nodes have very low posterior probabilities; we caution that no conclusions should be drawn from these branching events as they will be informed by the phylogenetic prior distribution rather than variable nucleotide sites (Figure 3A). Nevertheless, five clusters (denoted A-E) containing Guangdong sequences had posterior probability support of >80% (i.e. their sequences grouped monophyletically in over 80% of trees in the posterior sample; Figure 3B). These clusters are also observed in ML phylogeny (Figure 3A). Some included only sequences sampled in Guangdong (A, B), others included sequences sampled in other countries and provinces (C, D, E).

**Figure 3:**
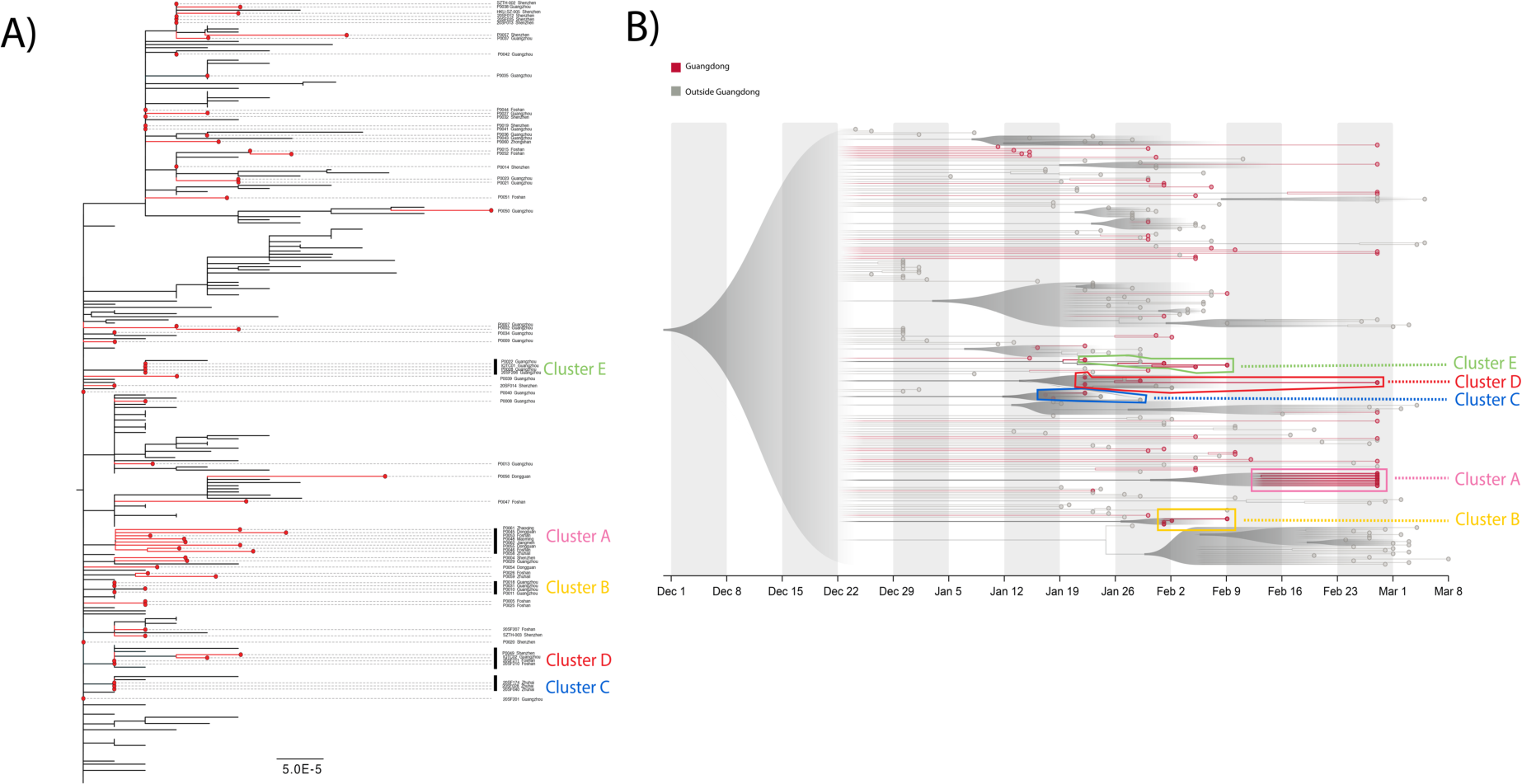
(A) Estimated maximum likelihood phylogeny of SARS-CoV-2 sequences from Guangdong (red branches and circles) and genomes from other countries and provinces (black). The tree is rooted using the reference strain MN908947.3. The scale bar is in units of expected nucleotide changes per site. Sampling locations within Guangdong are shown next to the sequence identifier. A phylogenetic bootstrap analysis was not performed due to the low number of phylogenetically informative sites and the number of missing bases (N) in the alignment. (B) Visualisation of the corresponding time-scaled maximum clade credibility tree. Sequences from Guangdong are in red and those from other locations in grey. The clusters (A-E) discussed in main text are highlighted with boxes. All nodes with posterior probabilities <0.5 have been collapsed into polytomies and their range of divergence dates are illustrated as shaded grey expanses.

From the molecular clock analysis, we were able to estimate the times of the most recent common ancestor (tMRCA) of clusteres A-E. We find that SARS-CoV-2 lineages were imported multiple times into Guangdong during the second half of January 2020 (Figure 4). Three clusters (C, D, E) have earlier tMRCAs that coincide with the start of the Guangdong epidemic and two (A, B) have later tMRCAs, around the time of the epidemic peak in the province (Figure 4). The average time between the tMRCA and the earliest sequence collection date in each cluster was approximately 10.5 days. The observed duration of each phylogenetic cluster (tMRCA to most recently sampled sequence) ranged from 13.8 (cluster B) to 45.5 (cluster D) days. The clusters with earlier tMRCAs contain more sequences sampled from travellers outside of China, possibly reflecting a decrease in air passenger travel from Guangdong after January 2020 (Flightradar 2020). The median tMRCA estimate of the COVID-19 pandemic was 29^th^ November 2019 (95% HPD 14^th^ November to 13^th^ December 2019), consistent with previous analyses (Rambaut 2020) (Figure 4).

**Figure 4:**
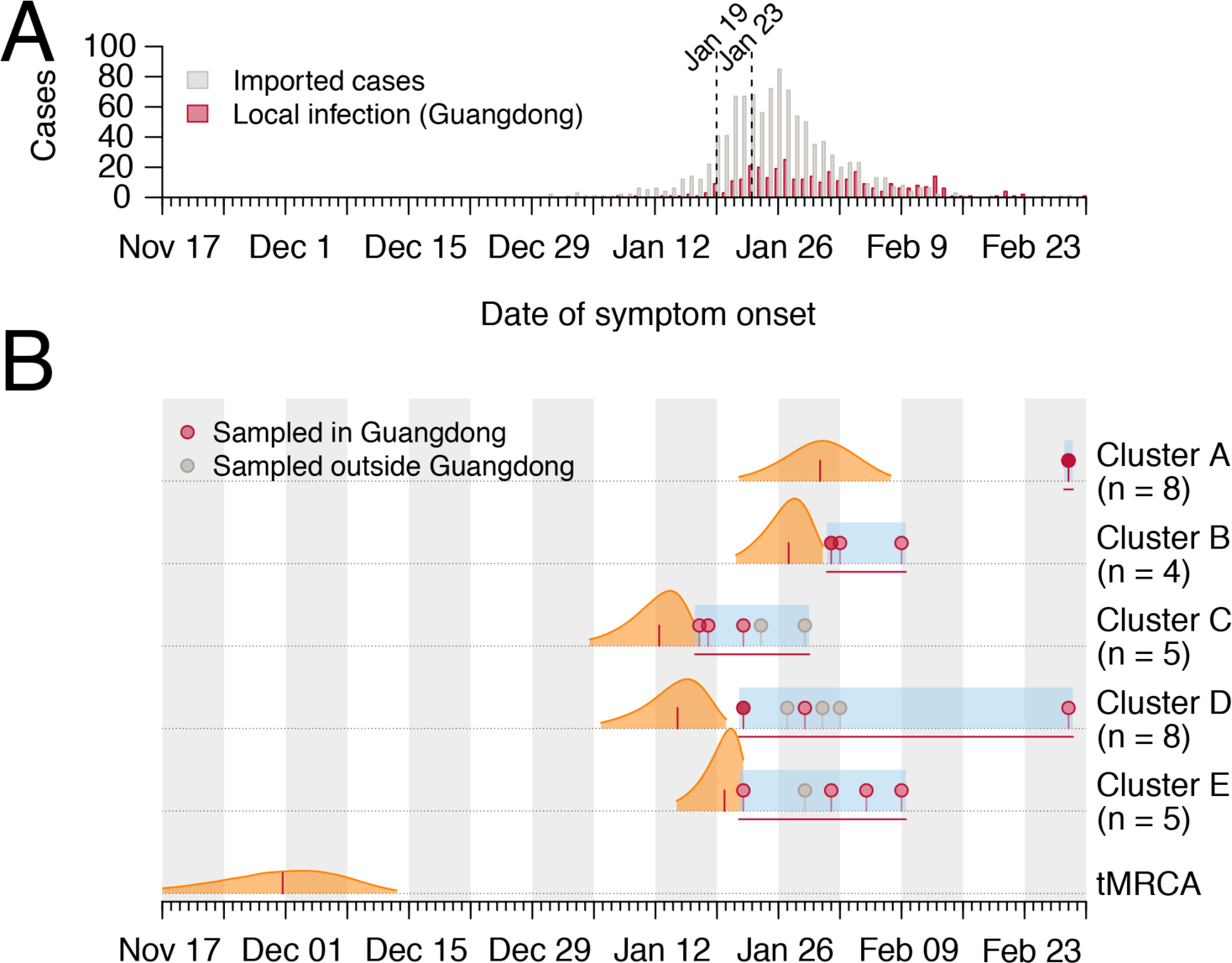
Molecular clock analysis of the five phylogenetic clusters of Guangdong sequences that were supported with posterior probabilities >80% in Bayesian phylogenetic analysis (A) Daily number of local and imported COVID-19 cases in Guangdong province, with the first reported case in Guangdong (Jan 19) and the shutdown of travel from Wuhan (Jan 23) indicated by dashed lines. (B) Posterior distributions of the tMRCAs of the five phylogenetic clusters (A-E) from the molecular clock analysis (Figure 3B). Distributions are truncated at the upper and lower limits of the 95% HPD intervals; the vertical red lines indicate median estimates. Blue shading and horizontal red lines indicate the sampling period over which genomes in each cluster were collected. Dots indicate the collection dates of genomes, coloured by sampling location (red = Guangdong, grey = other).

The apparent clusters of Guangdong sequences require careful interpretation, because of the relative undersampling of SARS-CoV-2 genomes from other Chinese provinces, including Hubei. Specifically, it is known that undersampling of regions with high incidence can lead to phylogenetic analyses underestimating the number of introductions into recipient locations, and overestimating the size and duration of transmission chains in those recipient locations (Grubaugh et al., 2017; Kraemer et al., 2018). For example, the largest Guangdong phylogenetic cluster comprises 8 sequences (denoted A in Figure 3,4), none of which are placed at the root of the cluster, and it is tempting to conclude that the whole cluster derived from community transmission within Guangdong. However, 6 of the 8 genomes in this cluster reported travel from Hubei and therefore the cluster in fact represents multiple SARS-CoV-2 introductions into Guangdong, with dates of symptom onset around or shortly after the shutdown of travel from Wuhan (Figure 4).

Our analyses of the genomic epidemiology of SARS-CoV-2 in Guangdong province indicate that, following the first COVID-19 case detected in early January, most infections were the result of virus importation from elsewhere, and that chains of local transmission were limited in size and duration. This suggests that the large-scale surveillance and intervention measures implemented in Guangdong were effective in interrupting community transmission in a densely populated urban region, ultimately containing the epidemic and limiting the potential for dissemination to other regions. However, vigilance is still required as the risk remains that SARS-CoV-2 transmission will reignite in Guangdong, following a recent increase in the number of COVID-19 cases imported to China from other countries.

The results also suggest that early phylogenetic analyses of the pandemic should be interpreted carefully. The number of mutations that define phylogenetic lineages are small (often one), and may be similar to the number of sequence differences arising from errors introduced during reverse transcription, PCR amplification, or sequencing. Bayesian estimates of divergence times (Rannala and Yang, 1996), such as the tMRCA of the pandemic, are based on aggregate numbers of mutations and are expected to be more robust. Further, the low and variable sampling of COVID-19 cases among different regions makes it challenging to evaluate phylogenetic clusters that comprise cases from a single region; although such clusters could indeed represent local transmission, our results show they can also include multiple introductions from a genomically-undersampled location. Therefore, as with all phylogenetic analyses, the SARS-CoV-2 genomes must be interpreted in the context of all available epidemiological information.

## Data Availability

All sequences data have been submitted to the public database (GISAID).

https://platform.gisaid.org/epi3/frontend

## Data Availability

The new sequences have been deposited in GISAID with accession IDs EPI_ISI_413850–413902.

## Acknowledgments & Funding

We gratefully acknowledge the efforts of local CDCs, hospitals and the third-party detection institutions in epidemiological investigations, sample collection, and detection. This work was supported by grants from Guangdong Provincial Novel Coronavirus Scientific and Technological Project (2020111107001), Science and Technology Planning Project of Guangdong (2018B020207006). OP, MUGK and LdP were supported by the Oxford Martin School. V.H. is supported by the Biotechnology and Biological Sciences Research Council (BBSRC) (grant number BB/M010996/1). NRF acknowledges funding from a Wellcome Trust and Royal Society Sir Henry Dale Fellowship (204311/Z/16/Z) and from a Medical Research Council and FAPESP award (MR/S0195/1). This work was supported by the Wellcome Trust ARTIC network. We would like to thank all the authors who have kindly deposited and shared genome data on GISAID. A table with genome sequence acknowledgments can be found in Supplementary Table S2.

## Methods

### Ethics

This study was approved by the institutional ethics committee of the Center for Disease Control and Prevention of Guangdong Province. Written consent was obtained from patients or their guardian(s) when samples were collected. Patients were informed about the surveillance before providing written consent, and data directly related to disease control were collected and anonymized for analysis.

### Sample collection, clinical surveillance and epidemiological data

After reports of hospitalized cases with undiagnosed, severe pneumonia on December 30^th^ 2019, enhanced surveillance was initiated in Guangdong Province to detect suspected infections, especially among cases with recent travel history to Hubei or other epidemic regions over the last 14 days. Suspected COVID-19 cases were screened by 31 designated hospitals, local CDCs in 21 prefecture cities, and 6 third-party detection institutions with commercial real-time reverse transcription PCR (RT-PCR) kits (China CDC, 2020). Further details of surveillance and case reporting are provided in Supplementary Text 1. A subset of positive samples was sent to Guangdong Provincial CDC for verification and further sequencing. All patients or their guardian(s) were informed about the molecular surveillance prior to providing written consent. Imported infections were defined when confirmed cases had travel history from Hubei or other epidemic regions and did not have close contact with local positive cases in 14 days preceding illness onset. The severity of the disease was classified into mild, moderate, severe, or critical type based on the Diagnosis and Treatment Scheme of SARS-CoV-2 released by the National Health Commission of China. Further details of clinical case definitions are provided in Supplementary Text 2. Demographic information, date of illness onset, and clinical outcomes of sequenced cases were collected from medical records. The exposure history for each case was obtained through an interview. Information regarding the demographic and geographic distribution of SARS-CoV-2 cases can be found at the website of Health Commission of Guangdong Province (http://wsjkw.gd.gov.cn/xxgzbdfk/yqtb/).

### Virus amplification and sequencing

Virus genomes were generated by two different approaches, (i) untargeted metagenomic sequencing on the BGI MGISEQ-2000 (n=63) and Illumina NextSeq (n=4) sequencing platforms, and (ii) using version 1 of the ARTIC COVID-19 multiplex PCR primers (https://artic.network/ncov-2019), followed by nanopore sequencing on an ONT MinION (n=45). We report only those genomes for which we were able to generate >50% genome coverage, and only report one genome per patient.

For metatranscriptomics, total RNAs were extracted from different types of samples by using QIAamp Viral RNA Mini Kit (Qiagen, Cat. No. 52904), followed by DNase treatment and purification with TURBO DNase (Invitrogen, Cat. No. AM2239) and Agencourt RNAClean XP beads (Beckman Cat. No. A63987). Both the concentration and the quality of all isolated RNA samples were measured and checked with the Agilent Bioanalyzer 2100 and Qubit. Libraries were prepared using the SMARTer Stranded Total RNA-Seq Kit v2 (Clontech, Cat. No. 634412) according to the manufacturer’s protocol starting with 10 ng total RNA. Briefly, purified RNA was firstly fragmented and converted to cDNA using reverse transcriptase. The ribosome cDNA was depleted by using ZapRv2 (mammalian-specific). The remaining cDNA was converted to double stranded DNA and subjected to end-repair, A-tailing, and adapter ligation. The constructed libraries were amplified using 9-16 PCR cycles. Sequencing of metatranscriptome libraries was conducted using Illumina NextSeq 550 SE 75 or BGI MGISEQ-2000 PE100 platforms.

For the multiplex PCR approach, we followed the general method of multiplex PCR as described in (https://artic.network/ncov-2019) (Quick et al., 2017). Briefly, the multiplex PCR was performed with two pooled primer mixture and the cDNA reverse transcribed with random primers was used as a template. After 35 rounds of amplification, the PCR products were collected and quantified, followed with end-repairing and barcoding ligation. Around 50 fmol of final library DNA was loaded onto the MinION. The nanopore sequencing platform takes less than 24 hours to obtain 10Gb of sequencing data, achieving between 0.3–0.6 million reads per sample. The ARTIC bioinformatics pipeline for COVID (https://artic.network/ncov-2019) was used to generate consensus sequences and call single nucleotide variants relative to the reference sequence.

To test the precision and threshold of the multiplex PCR and nanopore sequencing method, we undertook a serial dilution experiment. Viral RNA was extracted from a cell strain of SARS-CoV-2. To mimic clinical samples with different viral loads, we diluted this viral RNA with SARS-CoV-2 negative RNA extracted from nasopharyngeal swab specimens. Viral loads were estimated using RT-PCR with serial diluted plasmid as a standard. At each dilution level we performed multiplex PCR and nanopore sequencing and assembly as per the approach above, except that reads were assembled against the consensus genome obtained from the original sample using metagenomic sequencing. As expected, relative virus load, % genome coverage and average read depth decreased at higher dilutions. Genome coverage exceeded 75% for all except the final dilution (Table S3).

### Virus genome assembly

Reference-based assembly of the metagenomic raw data was performed as follows. Illumina adaptors were removed, and reads were filtered for quality (q30 threshold and read length >15nt) using Cutadapt 1.18 (Martin, 2011). The mapping of cleaned reads was performed against GenBank reference strain *MN908947*.*3* using Bowtie2 (Langmead and Salzberg, 2012). Consensus sequences were generated using samtools 1.2 (Li et al., 2009). Sites were called at depth>=3 if they matched the reference strain, or depth>=5 if they differed from the reference, otherwise sites were denoted N. Ambiguity nucleotide codes were used if (i) the minor variant is observed at >30% frequency and (ii) the minor variant is represented by 5 or more reads. Assembly of the nanopore raw data was performed using the ARTIC bioinformatic pipeline for COVID-19 with minimap2 (Li, 2018) and medaka (https://github.com/nanoporetech/medaka) for consensus sequence generation. For patient samples that were sequenced using both metagenomics and nanopore sequencing, we retained only the sequence with the highest genome coverage.

### Phylogenetic analysis

All available SARS-CoV-2 sequences (n=323) on GISAID (gisaid.org) on 13^th^ March 2020 were downloaded. Sequences from GISAID that were error-rich, those which represented multiple sequences from the same patient, and those without a date of sampling were removed. Finally, the dataset was reduced by only retaining the earliest and most recently sampled sequences from epidemiologically linked outbreaks (e.g. the Diamond Princess cruise ship). The resulting dataset of 250 sequences therefore represents the global diversity of the virus while minimizing the impact of sampling bias. Sequences were aligned using MAFFT v7.4 (Katoh and Standley, 2013) and manually inspected in Geneious v11.0.3 (https://www.geneious.com). The final alignment length was 29,923 nucleotides.

We used both the maximum likelihood (ML) and Bayesian coalescent methods to explore the phylogenetic structure of SARS-CoV-2. The ML phylogeny was estimated with PhyML (Guindon et al., 2010) using the HKY+Γ_4_ substitution model (Hasegawa et al., 1985) with gamma-distributed rate variation (Yang, 1994). Linear regression of root-to-tip genetic distance against sampling date indicated that the SARS-CoV-2 sequences evolve in clock-like manner (*r* = 0.539) (Figure S4). The Bayesian coalescent tree analysis was undertaken in the BEAST (Ayres et al., 2012; Suchard et al., 2018) framework, also using the HKY+Γ4 substitution model with gamma-distributed rate variation with an exponential population growth tree prior and a strict molecular clock. Taxon sets were defined and used to estimate the posterior probability of monophyly and the posterior distribution of the tMRCA of observed phylogenetic clusters A-E (Supplementary Table 1). Four independent chains were run for 100 million states and parameters and trees were sampled every 10,000 states. Upon completion, chains were combined using LogCombiner after removing 10% of states as burn-in and convergence was assessed with Tracer (Rambaut et al., 2018). The maximum clade credibility (MCC) tree was inferred from the Bayesian posterior tree distribution using TreeAnnotator, and visualised with figtreejs-react (https://github.com/jtmccr1/figtreejs-react@9874a5b). Code for Figure 3B is available at https://github.com/jtmccr1/tree-for-oli and a live version of the tree can be found at https://jtmccr1.github.io/tree-for-oli/. Monophyly and tMRCA (times to the most recent common ancestor) statistics were calculated for each taxon set from the posterior tree distribution.

## Notes

### Competing Interest Statement

The authors have declared no competing interest.

